# SARS-CoV-2 infection in households with and without young children: Nationwide cohort study

**DOI:** 10.1101/2021.02.28.21250921

**Authors:** Anders Husby, Giulia Corn, Tyra Grove Krause

## Abstract

**Background:** Infections with seasonally spreading human coronaviruses (HCoVs) are common among young children during winter months in the northern hemisphere, with immunological response lasting around a year. However, it is not clear whether recent household exposure to HCoVs reduces the risk of SARS-CoV-2 infection.

**Methods:** In a nationwide cohort study we followed all adults in Denmark aged 18 to 60 years from February 27 to November 15, 2020. Hazard ratios of SARS-CoV-2 infection by number of young children aged ten months to five years living in the household were estimated using Cox regression adjusted for adult age, gender, and other potential confounders. In sensitivity analyses we investigated the effect of age of children in the household, number of children living outside of the household, and number of other adult household-members.

**Results:** Among 449,915 adults in Denmark living in households with young children, 5,761 were tested positive for SARS-CoV-2, while among 2,629,821 adults without young children in their household, 33,788 were tested positive for SARS-CoV-2 (adjusted hazard ratio, 1.05; 95% confidence interval, 1.02 to 1.09). Sensitivity analyses of age of children in the household, number of children living outside of the household, and number of additional adult household members found increasing number of children, and especially increasing number of older children, to substantially increase the risk of SARS-CoV-2 infection.

**Conclusions:** Living in a household with young children was not associated with decreased risk of SARS-CoV-2 infection, thereby suggesting no strong preventive effect of recent exposure to HCoVs against SARS-CoV-2 infection.

## BACKGROUND

Infection with the novel coronavirus SARS-CoV-2 only has limited treatment options(1–3) and immunity against the virus is globally very limited(4). Nevertheless, it is suggested that recent exposure to seasonally spreading human coronaviruses (HCoVs, e.g. OC43 and NL63), which is particularly prevalent among young children during winter months(5), might result in protection from SARS-CoV-2 infection(6,7). Furthermore, studies report serological evidence of pre-existing SARS-CoV-2 binding antibodies(8) and T-cell immunity(9–11) in individuals not infected with SARS-CoV-2, thought to arise from previous HCoV infection. However, humoral immunity against HCoVs is short-lived and immunological cross-reactivity of HCoVs against SARS-CoV-2 may not be sterilizing(12). It is therefore unknown if, and to which degree, recent exposure to HCoVs reduces SARS-CoV-2 infection risk or disease severity.

Taking advantage of complete individual-level data on inhabitants of all households in Denmark and nationwide information on all laboratory-confirmed SARS-CoV-2 infections and hospitalizations, we explored the role of exposure to HCoVs on risk of SARS-CoV-2 infection and hospitalization, using living in a household with young children as a proxy for recent HCoV exposure.

## METHODS

### Materials

The Danish Civil Registration System provides demographic information on the Danish population, in addition to information on household members and number of offspring(13). Information on PCR-tests for SARS-CoV-2 is available through MiBA, the Danish Microbiology Database, which includes all microbiological test results from public laboratories in Denmark(14). Denmark has one of the highest SARS-CoV-2 PCR testing capacities in Europe, with free tests easily accessible and offered to all inhabitants through the public healthcare system(15). Information on all hospitalizations in Denmark is available through the Danish National Patient Registry(16).

### Study population

All adults aged 18 to 60 years living in Denmark on January 1, 2020, with known address were included in the study cohort. In addition, we constructed a cohort of SARS-CoV-2-test-positive individuals who were followed up for hospitalization until 30 days after positive test.

We excluded adults living in households with seven or more individuals (only 1.7 % of the population in Denmark), to avoid inclusion of households consisting of multiple families (e.g. collective housing communities living on the same address).

### Exposure

The primary exposure was defined as living, per January 1, 2020, in a household with one or more children aged ten months to five years. In this age span children are usually enrolled in childcare institutions and seroconvert against seasonal coronaviruses(17–19). The exposure is therefore a proxy of recent close contact with a child infected with HCoVs, as used previously(7). To ensure validity of a close relationship between adult and children, only individuals who were legal parents to the child or children were included as exposed in the primary exposure analysis. In the sensitivity analysis we examined other types of co-residence.

### Study covariates

In addition to age and gender, we adjusted for urbanicity by grouping the 98 Danish municipalities into ten groups based on population density, from most rural to most urban. Additionally, we adjusted for ethnicity, as an earlier report found higher incidence of SARS-CoV-2 infection among immigrant groups in Denmark(20). Furthermore, we adjusted for the following comorbidities: asthma, chronic pulmonary disease (incl. COPD), cardiovascular disease, diabetes mellitus, inflammatory bowel disease, malignancy, and renal failure (see definition of comorbidities in Table S1).

### Outcomes

The main outcome was a positive SARS-CoV-2 test from February 27 (date of first positive SARS-CoV-2 test in Denmark). As secondary outcome we investigated risk of hospitalization 30 days following a positive SARS-CoV-2 test. See supplementary Figure S1 for overview of daily SARS-CoV-2 test positivity rate, cases, and hospital admissions during the study period.

### Statistical analysis

Hazard ratios for SARS-CoV-2 infection by household type were estimated using Cox regression with calendar period as the underline time scale, adjusting for gender and age at entry and using a robust variance structure to account for the correlation between members of the same household. Cohort members were followed from February 27, 2020, to outcome of interest, death, emigration, or November 15, 2020, whichever came first. In a secondary analysis, we investigated the 30-day hazard ratio of hospitalization among individuals testing positive for SARS-CoV-2 by household type.

In sensitivity analyses we investigated interaction with adult age, gender, and period of testing (February 27-March 26 (before lockdown), March 27-April 28 (lockdown), April 29-June 30 (early reopening), July 1-November 15 (late reopening/resurgence of community transmission). Furthermore, we investigated age span of the exposure definition, different types of households based on co-residence and legal parenthood, and different types of households with and without co-residence of young and older children. In addition, we estimated the effect of number of adults in the household.

To investigate whether adults living with young children were tested more compared with other adults, we compared the incidence rate ratio (IRR) of SARS-CoV-2 PCR testing within the latest 60 days among the two groups using Poisson regression. All tests until the first positive test, if any, were included in the model as outcome.

Finally, we estimated the hazard ratio of SARS-CoV-2 infection according to age and number of all household children (age <18 years) relative to only-children aged 6 years. Each adult contributed with a number of observations equal to their number of children. The model included a restricted cubic spline term with 4 knots (located at the 5th, 35th, 65th, and 95th-percentile of the age distribution) for child age, a 3-level variable for number of children (one, two, or three or more children) and a robust variance structure and was adjusted for the covariates included in the previous analyses. The Bayesian Information Criterion was used to choose between a model with an interaction term for child age and number of household children and a model with additive effect.

## RESULTS

In our cohort of 3,079,736 adults living in Denmark aged 18 to 60 years, 449,915 (14.6%) lived in households with young children aged ten months to five years, while 2,629,821 (85.4%) lived in households without young children (Table 1). Adults living with young children were, on average, younger (median age 35 versus 42 years), and more often female (54% versus 49%) than adults not living with young children. For both groups, the most predominant household type consisted of two adults (86% among adults living with young children and 49% among adults not living young children). For the group of adults in households with young children, 76%, 23%, and 1% lived with one, two, or three or more young children, respectively. For adults in households with any children below 18 years, 35%, 48%, and 18% lived with one, two, or three or more children, respectively. Medical comorbidities, as defined from nationwide hospital diagnostic codes, were more common among individuals not living in households with young children, except for inflammatory bowel disease and renal failure.

**Table 1.**
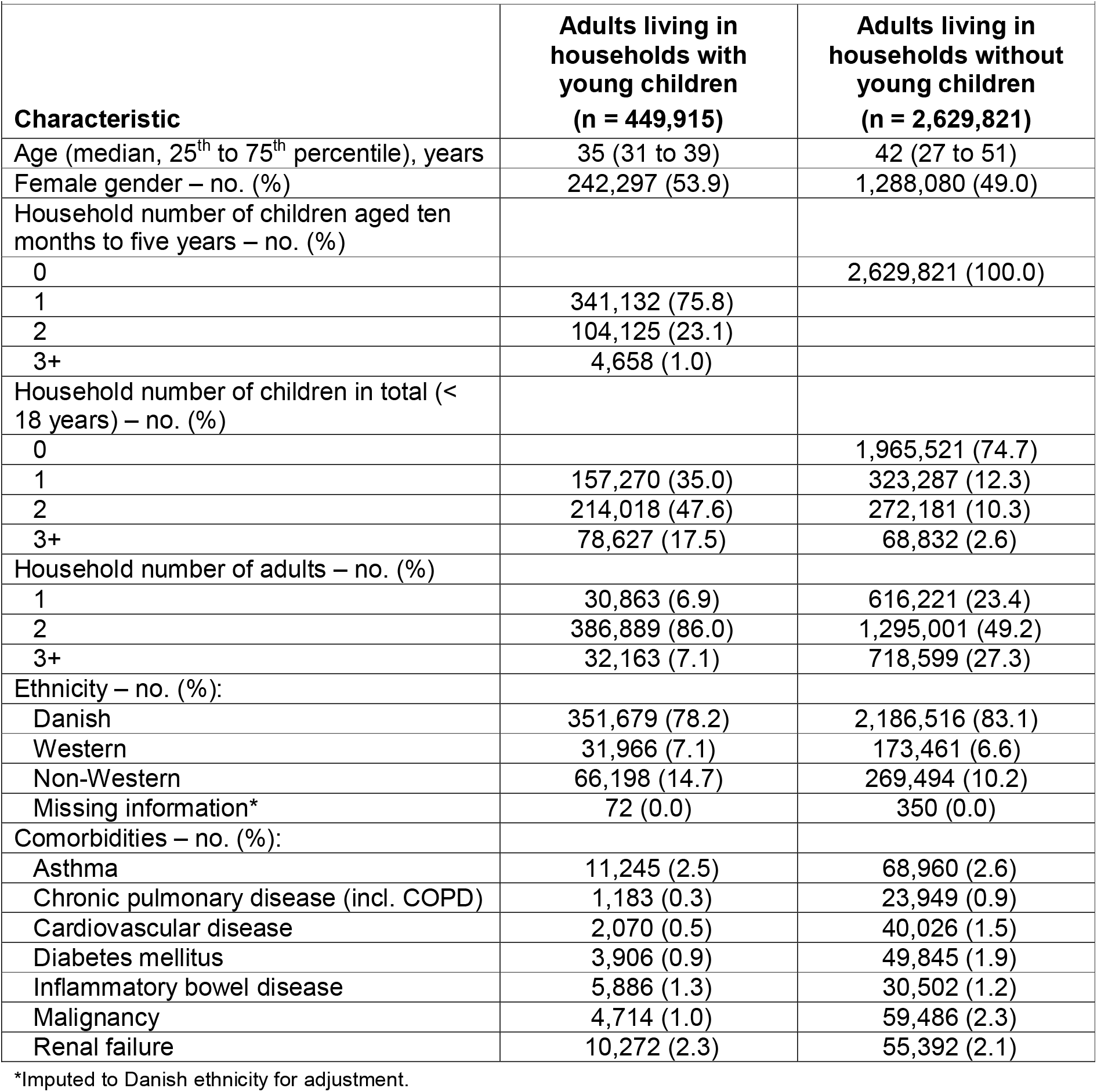
Baseline characteristics (age, gender, number of children, number of adults in the household, and comorbidities) of the cohort of 3,079,736 adults by household type.

| **Characteristic** | **Adults living in households with young children**<br>**(n = 449,915)** | **Adults living in households without young children**<br>**(n = 2,629,821)** |
| --- | --- | --- |
| Age (median, 25<sup>th</sup> to 75<sup>th</sup> percentile), years | 35 (31 to 39) | 42 (27 to 51) |
| Female gender – no. (%) | 242,297 (53.9) | 1,288,080 (49.0) |
| Household number of children aged ten months to five years – no. (%) |  |  |
| 0 |  | 2,629,821 (100.0) |
| 1 | 341,132 (75.8) |  |
| 2 | 104,125 (23.1) |  |
| 3+ | 4,658 (1.0) |  |
| Household number of children in total (< 18 years) – no. (%) |  |  |
| 0 |  | 1,965,521 (74.7) |
| 1 | 157,270 (35.0) | 323,287 (12.3) |
| 2 | 214,018 (47.6) | 272,181 (10.3) |
| 3+ | 78,627 (17.5) | 68,832 (2.6) |
| Household number of adults – no. (%) |  |  |
| 1 | 30,863 (6.9) | 616,221 (23.4) |
| 2 | 386,889 (86.0) | 1,295,001 (49.2) |
| 3+ | 32,163 (7.1) | 718,599 (27.3) |
| Ethnicity – no. (%): |  |  |
| Danish | 351,679 (78.2) | 2,186,516 (83.1) |
| Western | 31,966 (7.1) | 173,461 (6.6) |
| Non-Western | 66,198 (14.7) | 269,494 (10.2) |
| Missing information* | 72 (0.0) | 350 (0.0) |
| Comorbidities – no. (%): |  |  |
| Asthma | 11,245 (2.5) | 68,960 (2.6) |
| Chronic pulmonary disease (incl. COPD) | 1,183 (0.3) | 23,949 (0.9) |
| Cardiovascular disease | 2,070 (0.5) | 40,026 (1.5) |
| Diabetes mellitus | 3,906 (0.9) | 49,845 (1.9) |
| Inflammatory bowel disease | 5,886 (1.3) | 30,502 (1.2) |
| Malignancy | 4,714 (1.0) | 59,486 (2.3) |
| Renal failure | 10,272 (2.3) | 55,392 (2.1) |

\*Imputed to Danish ethnicity for adjustment.

When investigating risk of SARS-CoV-2 infection in adults living in households with young children compared with adults living in households without young children, we found overall an adjusted hazard ratio of 1.05 (95% CI, 1.02 to 1.09) of SARS-CoV-2 infection (Table 2). When stratifying by number of young children in the household we found an adjusted hazard ratio of 1.03 (95% CI, 0.99 to 1.07), 1.13 (95% CI, 1.05 to 1.21), 1.42 (95% CI, 1.09 to 1.84), of SARS-CoV-2 infection for living in a household with one, two, or three or more children, respectively, compared with individuals in households with no young children (P = 0.002 for trend).

**Table 2.** Relative risk of SARS-CoV-2 infection in adults by household type and number of young children.

|  |  |  |  |
| --- | --- | --- | --- |
| **Household type** | **Adult cases / Adults in total** | **Relative risk of SARS-CoV-2 infection** |  |
|  |  | *hazard ratio (95% CI)* |  |
|  |  | **Crude\*** | **Adjusted<sup>†</sup>** |
| Household without young children | 33,788 / 2,629,821 | 1 (ref.) | 1 (ref.) |
| Household with young children (any) | 5,761 / 449,915 | 1.04 (1.00-1.08) | 1.05 (1.02-1.09) |
| 1 | 4,322 / 341,132 | 1.03 (0.99-1.07) | 1.03 (0.99-1.07) |
| 2 | 1,358 / 104,125 | 1.07 (1.00-1.15) | 1.13 (1.05-1.21) |
| 3+ | 81 / 4,658 | 1.43 (1.10-1.85) | 1.42 (1.09-1.84) |

\*Crude: Age and gender adjusted only.
<sup>†</sup>Adjusted: Furthermore adjusted for urbanicity, ethnicity, and comorbidities.

Investigating the risk of SARS-CoV-2 hospitalization among SARS-CoV-2-positive adults by household status, we found a non-significant decreased risk of SARS-CoV-2 hospitalization among adults living in households with any number of young children compared with adults living without young children in the household (adjusted hazard ratio 0.84, 95% CI, 0.69 to 1.03) (Table 3). When stratifying by number of young children in the household we found an adjusted hazard ratio of 0.87 (95% CI, 0.70 to 1.09) and 0.75 (95% CI, 0.52 to 1.09) of SARS-CoV-2 hospitalization for adults living with one, or two or more children, respectively, compared with adults in households with no young children (P = 0.47 for trend).

**Table 3.** Relative risk of SARS-CoV-2 hospitalization in adults by household type and number of young children, among SARS-CoV-2-positive individuals.

| Household type | Adults hospitalized /<br>SARS-CoV-2 positive<br>adults in total | Relative risk of SARS-CoV-2 hospitalization<br>*hazard ratio (95% CI)* |  |
| --- | --- | --- | --- |
|  |  | Crude* | Adjusted† |
| Household without young children | 1,130 / 18,542 | 1 (ref.) | 1 (ref.) |
| Household with young children (any) | 146 / 3,093 | 0.86 (0.70-1.05) | 0.84 (0.69-1.03) |
| 1 | 113 / 2,278 | 0.88 (0.71-1.10) | 0.87 (0.70-1.09) |
| 2+ | 33 / 815 | 0.77 (0.53-1.12) | 0.75 (0.52-1.09) |

\*Crude: Age and gender adjusted only.
†Adjusted: Furthermore adjusted for urbanicity, ethnicity, and comorbidities.

Further, we explored the role of adult age, gender, and time-period of testing (Table S2), and found the relative SARS-CoV-2 infection risk to be noticeably higher among adults aged 30-39 years sharing a household with young children (adjusted hazard ratio 1.14 (95% CI, 1.08 to 1.19)) in comparison with other age strata, and especially high during early reopening of society following lockdown (April 29-June 30, 2020), adjusted hazard ratio of 1.26 (95% CI, 1.11 to 1.44). In addition, we found evidence of a significant, but small, gender effect, with higher risk of SARS-CoV-2 infection among men compared with women (P = 0.04 for interaction). Scrutinizing effects of age-span definition of young children, household definition of children, presence of older and younger children in the household (Table S3-S6), we found strong evidence of increased SARS-CoV-2 infection risk among adults with older children in the household, regardless of the presence of young children in the household (adjusted hazard ratio of 1.31 (95% CI, 1.24 to 1.37)) or not (adjusted hazard ratio of 1.31 (95% CI, 1.27 to 1.35)). In households with only young children, a clear pattern persisted of increased risk of infection by increasing number of young children. Different definitions of age span of young children and number of children in the household (i.e. inclusion or exclusion of non-legal children) did not markedly alter the effect of young children on SARS-CoV-2 infection risk. Additionally, we examined the role of number of adults in the household (Table S7). When stratifying by number of household adults, we found a heterogeneous pattern, where in households with two adults, there was no increased relative risk of SARS-CoV-2 infection in households with young children, while we observed an increased relative risk in households with one or three or more adults. Finally, as a last sensitivity analysis, we investigated SARS-CoV-2 testing intensity by household type (Table S8). We found a 9% increased testing rate in households with young children compared with households without young children. However, adjusting for testing intensity had minimal impact on our main finding, with an adjusted hazard ratio of 1.05 (95% CI, 1.01 to 1.09) for any young child when adjusted for tests within the latest 60 days.

To illustrate relative effects of child age, we plotted cubic splines of SARS-CoV-2 infection risk in adults living in households with children by child age and total number of children in the household relative to having one child aged six years (Figure 1). Overall, higher child age and higher number of children was associated with an additive increased relative risk of infection.

**Figure 1.**
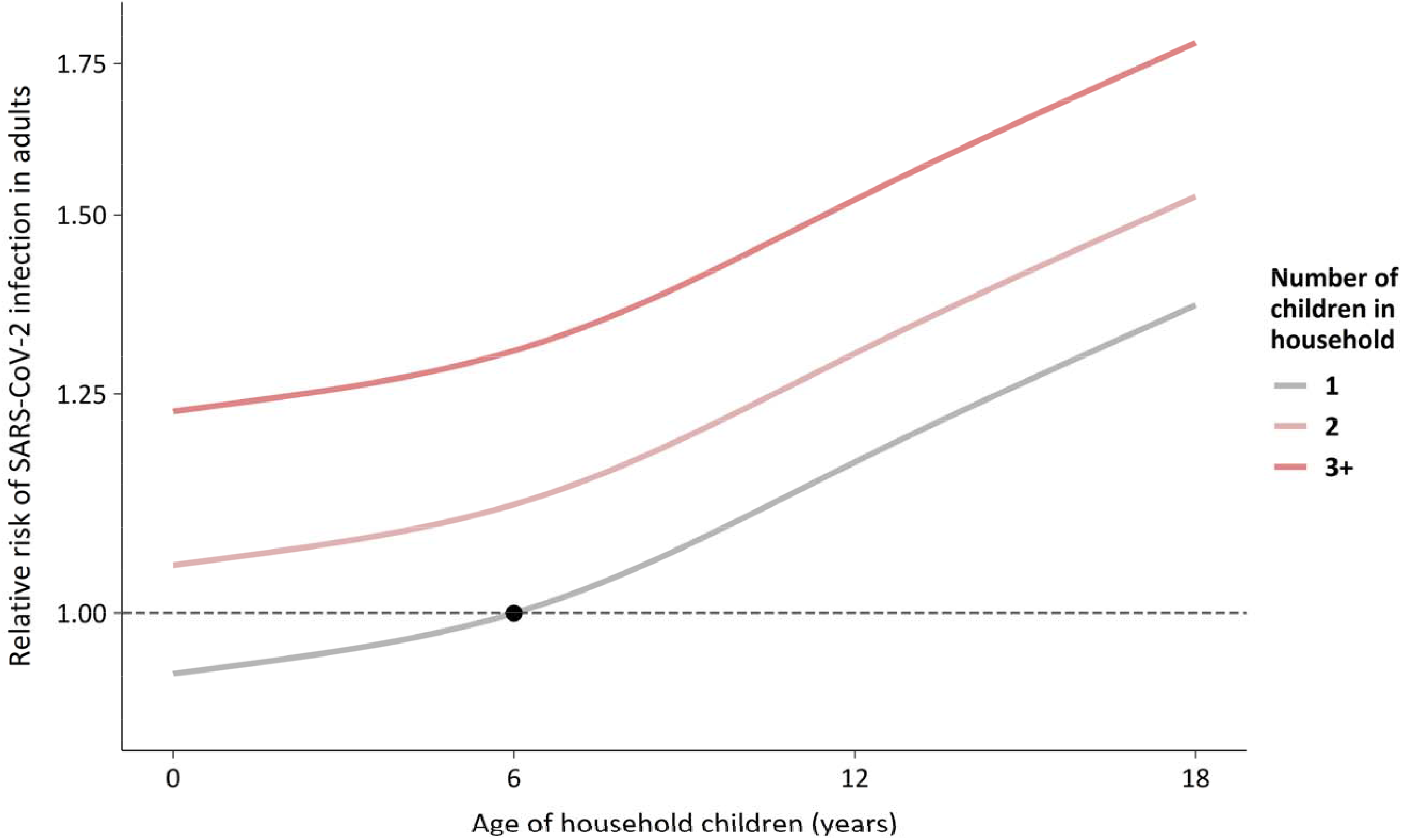
Relative risk of SARS-CoV-2 infection in adults living in households with children by child age and total number of children in the household relative to having one child aged six years. Adjusted for age, gender, urbanicity, ethnicity, and comorbidities.

## DISCUSSION

In a nationwide cohort study of all adults in Denmark aged 18 to 60 years, living in households with young children was associated with a small, but significant, increased risk of SARS-CoV-2 infection, compared with adults not living in households with young children, adjusting for potential confounders. The risk of infection was amplified with increasing number of young children living in the household, but the overall association was attenuated when excluding households with older children. Overall, the results suggest no reduced SARS-CoV-2 infection risk in individuals with recent exposure to HCoVs.

Our study is, to our knowledge, the first study of household characteristics and SARS-CoV-2 infection risk enrolling an entire population of more than three million individuals. Furthermore, the study likely captures the vast majority of SARS-CoV-2 infections in Denmark during most of the study period, as Denmark has one of the highest PCR testing rates in Europe(15). This is reflected in the low positivity rate of SARS-CoV-2 tests of 2% from late-April to the end of follow-up in mid-November 2020 (Figure S1).

Our study has some limitations. We did not have serological measurements of B-cell or T-cell immunity to HCoVs, which could offer direct biological evidence of preexisting immunity against SARS-CoV-2 following prior exposure to HCoVs, but co-habitation with young children has been considered a reasonable proxy for recent exposure to HCoVs previously(7). Furthermore, we did not include information on SARS-CoV-2 testing of young children. However, young children are often asymptomatic or present with mild symptoms of infection(21), thus inclusion of this information would introduce undesirable health-seeking behavior bias into our analyses.

Our findings show that having young children in one’s household was associated with a slightly increased risk of SARS-CoV-2 infection. The association could be a result of a higher number of social contacts among adults living in households with young children (e.g. more adults in the household, contact to daycare facilities, or close contact to parents of playmates) or due to infection brought into the household by the young children. Nevertheless, when we stratified by number of adults in the household we found no indication that an increased number of adults in the household was the driving force behind increased risk of SARS-CoV-2 infection. However, we found a heterogeneous pattern according to which the lowest relative risk of infection was in households with two adults, indicating that social circumstances, and not the household number of adults per se, is the most important factor in determining household infection risk. Furthermore, our analyses also show an increasing infection risk with increasing number of young children and increased relative risk of infection during early reopening in Denmark, where daycare facilities were among the first places to reopen, supporting that young children to some degree transmit SARS-CoV-2 in the household. Still, compared with older children, the transmission from young children within the household is relatively small (Figure 1). It is therefore imperative to weigh the benefits of young children attending daycare facilities and having playdates against the relatively small risk of SARS-CoV-2 transmission.

Our findings are in line with findings from serological and epidemiological studies that suggest no sterilizing cross-reactive immunity of recent exposure to HCoVs(5,8). As opposed to previous studies, we were also able to investigate the severity of SARS-CoV-2 infection in a population-setting, utilizing information on hospital admissions of all SARS-CoV-2 positive cases. We did not find statistical significant reduced risk of hospitalization among individuals living with young children compared with individuals living without. Nevertheless, the analyses had limited power due to the few hospitalizations of adults with young children, therefore, findings from other countries with similar data would be of value to explore a potential effect of HCoVs exposure on SARS-CoV-2 infection severity.

The newly emerging B.1.1.7 variant of SARS-CoV-2, was initially reported to be relatively more infectious in individuals below 20 years of age(22), but was very rare in Denmark during the study period and cannot explain our findings. However, given the surging spread of B.1.1.7, our study provides a framework for future population scale studies of transmission between children and their parents of this and other SARS-CoV-2 variants.

In summary, we found no evidence of a reduced risk of SARS-CoV-2 infection in adults living with young children. On the contrary, we found a significant, slightly increased risk of SARS-CoV-2 infection, which, however, was attenuated when taking account of older children in the same household. Our study thereby suggests no protection against SARS-CoV-2 infection from recent exposure to HCoVs.

## Supporting information

STROBE checklist

## Data Availability

The data used in the study can be obtained by first submitting a research protocol to the Danish Data Protection Agency (Datatilsynet) and once permission has been received, by applying the Ministry of Healths Research Service (Forskerservice) for access to the data. The data do not belong to the authors and they are not permitted to share them, except in aggregate form.

## Notes

### Competing Interest Statement

The authors have declared no competing interest.

### Funding Statement

No funding for this project.

### Author Declarations

Data availability statement: The datasets analyzed in the study are located in the Danish national COVID-19 surveillance system database at Statens Serum Institut, and the data are becoming or are already available for research upon reasonable request and with permission from the Danish Data Protection Agency and Danish Health and Medicines Authority. Ethics: The study was conducted on administrative register data. According to Danish law, ethics approval is exempt for such research, and the Danish Data Protection Agency, which is a dedicated ethics and legal oversight body, thus waives ethical approval for our study of administrative register data, where individual participants are not contacted and only aggregate results are presented. The study is therefore fully compliant with all legal and ethical requirements and there are no further processes available regarding such studies.

