## Supplementary appendix for "SARS-CoV-2 infection in households with and without young children: Nationwide cohort study"

### Supplementary Figures

**Figure S1.** SARS-CoV-2 epidemic in Denmark 2020. Daily SARS-CoV-2 test positivity rate (A), cases (B), and hospital admissions (C) up to November 15, 2020.

A

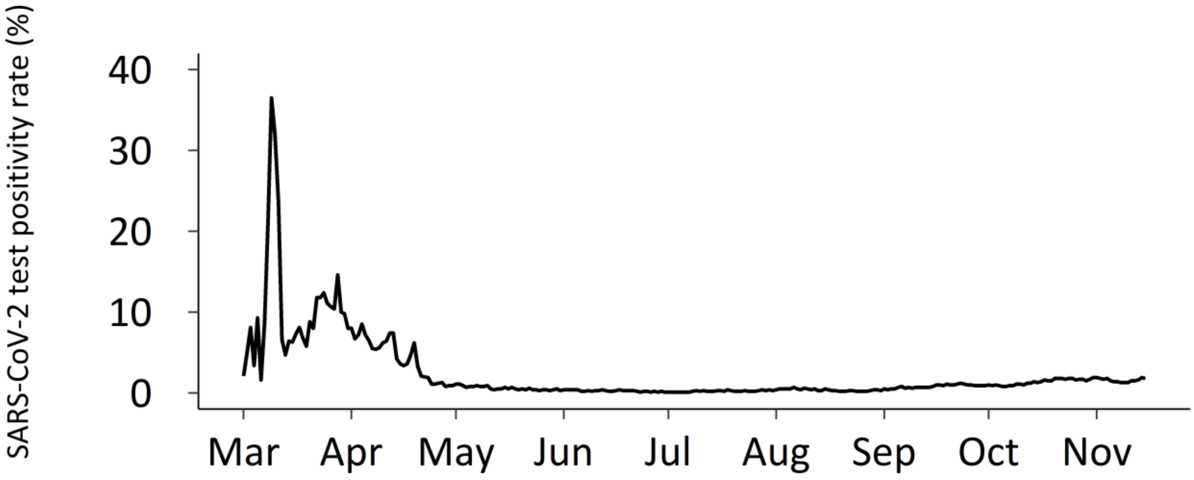

B

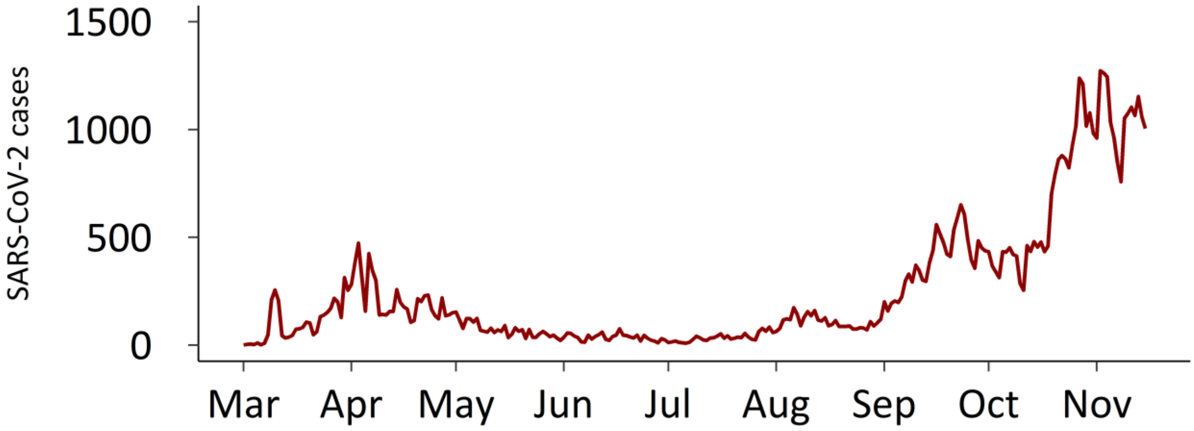

C

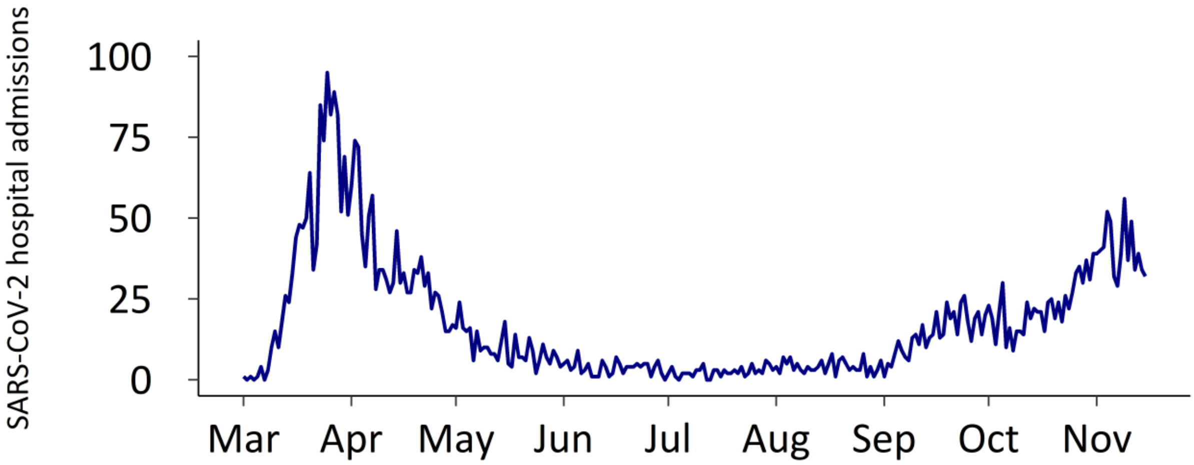

### Supplementary Tables

**Table S1.** Diagnostic codes used for ascertainment of comorbidities.

| *Comorbidity* | **ICD-10 codes** |
|---|---|
| Asthma | J45-J46 |
| Chronic pulmonary disease (incl. COPD) | J40-J44, J47, J60–J67, J68.4, J70.1, J70.3, J84.1, J92.0, J96.1, J98.2, J98.3 |
| Cardiovascular disease | I11.0, I13.0, I13.2, I21-I23, I48, I50x, |
| Diabetes mellitus | E10.0, E10.1, E10.9, E11.0, E11.1, E11.9 |
| Inflammatory bowel disease | K50x, K51x |
| Malignancy | C00–C75 |
| Renal failure | I12, I13, N00–N05, N07, N11, N14, N17–N19, Q61, N30.0 |

**Table S2.** Relative risk of SARS-CoV-2 infection in adults by household type and number of young children, by adult age, gender, and time period.

| | Adult cases with young children / Adults in total | Adult cases without young children / Adults in total | Relative risk of SARS-CoV-2 infection *hazard ratio (95% CI)* | |
|---|---|---|---|---|
| | | | Crude* | Adjusted† |
| **Age (years)‡** | | | | |
| 18-29 | 984 / 75,045 | 12,500 / 810,836 | 0.92 (0.85,0.99) | 1.01 (0.93,1.09) |
| 30-39 | 3,526 / 278,125 | 4,503 / 386,277 | 1.08 (1.03,1.13) | 1.14 (1.08,1.19) |
| 40-59 | 1,251 / 96,745 | 16,785 / 1,432,708 | 1.09 (1.02,1.17) | 0.95 (0.89,1.02) |
| **Gender§** | | | | |
| Female | 3,161 / 242,297 | 17,277 / 1,288,080 | 1.01 (0.97,1.06) | 1.03 (0.99,1.07) |
| Male | 2,600 / 207,618 | 16,511 / 1,341,741 | 1.08 (1.03,1.13) | 1.08 (1.03,1.13) |
| **Time period¶** | | | | |
| February 27-March 26 (pre-lockdown) | 166 / 449,915 | 1,101 / 2,629,821 | 0.92 (0.78,1.10) | 0.93 (0.79,1.11) |
| March 27-April 28 (lockdown) | 684 / 449,537 | 3,876 / 2,626,103 | 1.08 (0.99,1.19) | 1.09 (1.00,1.20) |
| April 29-June 30 (early reopening) | 366 / 448,746 | 1,799 / 2,620,713 | 1.25 (1.09,1.42) | 1.26 (1.11,1.44) |
| July 1-November 15 (late reopening) | 4,545 / 448,004 | 27,012 / 2,614,231 | 1.03 (0.99,1.07) | 1.04 (1.00,1.08) |

*Crude: Age and gender adjusted only.

†Adjusted: Furthermore adjusted for urbanicity, ethnicity, and comorbidities.

‡ P-value for interaction <0.0001 in adjusted analysis.

§ P-value for interaction = 0.04 in adjusted analysis.

¶ P-value for interaction = 0.02 in adjusted analysis.

| **Table S3.** Relative risk of SARS-CoV-2 infection in adults by household type and number of young children, by age span definition. | | | | |
|---|---|---|---|---|
| **Age span definition** | **Adult cases with young children / Adults in total** | **Adult cases without young children / Adults in total** | **Relative risk of SARS-CoV-2 infection** *hazard ratio (95% CI)* | |
| | | | **Crude*** | **Adjusted†** |
| 1-3 years | 3,908 / 309,685 | 35,641 / 2,770,051 | 1.01 (0.97-1.06) | 1.03 (0.99-1.08) |
| 10 months – 5 years | 5,761 / 449,915 | 33,788 / 2,629,821 | 1.04 (1.00-1.08) | 1.05 (1.02-1.09) |
| 0-9 years | 8,499 / 658,639 | 31,050 / 2,421,097 | 1.06 (1.03-1.10) | 1.08 (1.04-1.11) |

*Crude: Age and gender adjusted only.

†Adjusted: Furthermore adjusted for urbanicity, ethnicity, and comorbidities.

| **Table S4.** Relative risk of SARS-CoV-2 infection in adults by household type and number of young children, by exposure with all young children in household and out-of-household. | | | | |
|---|---|---|---|---|
| | | | **Relative risk of SARS-CoV-2 infection** *hazard ratio (95% CI)* | |
| **Household children definition** | **Adult cases with young children / Adults in total** | **Adult cases without young children / Adults in total** | **Crude*** | **Adjusted†** |
| All legally parented young children | 6,129 / 480,733 | 33,420 / 2,599,003 | 1.04 (1.00,1.08) | 1.05 (1.01,1.09) |
| All household co-living young children | 6,234 / 476,038 | 33,315 / 2,603,698 | 1.07 (1.03,1.11) | 1.05 (1.02,1.09) |
| Only legally parented and co-living household young children | 5,761 / 449,915 | 33,788 / 2,629,821 | 1.04 (1.00,1.08) | 1.05 (1.02,1.09) |

*Crude: Age and gender adjusted only.

†Adjusted: Furthermore adjusted for urbanicity, ethnicity, and comorbidities.

| **Table S5.** Relative risk of SARS-CoV-2 infection in adults by household type and number of young and older children in household. | | | |
|---|---|---|---|
| | | **Relative risk of SARS-CoV-2 infection** *hazard ratio (95% CI)* | |
| **Household type** | **Adult cases / Adult in total** | **Crude*** | **Adjusted†** |
| Household without any children | 24,571 / 1,965,521 | 1 (ref.) | 1 (ref.) |
| Household with young children and without older children (≥ 6 years) | 2,822 / 242,259 | 1.01 (0.96,1.06) | 1.04 (0.99,1.10) |
| Household with young children and with older children (≥ 6 years) | 2,939 / 207,656 | 1.30 (1.23,1.36) | 1.31 (1.24,1.37) |
| Household without young children and with older children (≥ 6 years) | 9,217 / 664,300 | 1.28 (1.24,1.32) | 1.31 (1.27,1.35) |

Crude: Age and gender adjusted only.

Adjusted: Furthermore adjusted for urbanicity, ethnicity, and comorbidities.

| **Table S6.** Relative risk of SARS-CoV-2 infection in adults living in household with young children, and not older children, by number of young children. | | | |
|---|---|---|---|
| **Number of young children** | **Adult cases / Adults in total** | **Relative risk of SARS-CoV-2 infection** *hazard ratio (95% CI)* | |
| | | **Crude*** | **Adjusted†** |
| 1 | 1,768 / 157,270 | 1 (ref.) | 1 (ref.) |
| 2 | 990 / 81,064 | 1.12 (1.02,1.23) | 1.22 (1.11,1.34) |
| 3+ | 64 / 3,925 | 1.50 (1.11,2.03) | 1.54 (1.14,2.08) |

*Crude: Age and gender adjusted only.

†Adjusted: Furthermore adjusted for urbanicity, ethnicity, and comorbidities.

| **Table S7.** Relative risk of SARS-CoV-2 infection in adults by household type and by number of adults in household. | | | | |
|---|---|---|---|---|
| **Number of adults in household** | **Adult cases with young children / Adults in total** | **Adult cases without young children / Adults in total** | **Relative risk of SARS-CoV-2 infection** *hazard ratio (95% CI)* | |
| | | | **Crude*** | **Adjusted†** |
| 1 | 393 / 30,863 | 5,842 / 616,221 | 1.26 (1.14,1.40) | 1.22 (1.10,1.35) |
| 2 | 4,689 / 386,889 | 16,014 / 1,295,001 | 0.98 (0.94,1.02) | 1.00 (0.96,1.04) |
| 3+ | 670 / 32,163 | 11,932 / 718,599 | 1.28 (1.16,1.41) | 1.12 (1.02,1.24) |

*Crude: Age and gender adjusted only.

†Adjusted: Furthermore adjusted for urbanicity, ethnicity, and comorbidities.

| **Table S8.** Relative risk (incidence rate ratio) of test for SARS-CoV-2 in adults by household type. | | | | | | |
|---|---|---|---|---|---|---|
| **Household type** | **Adults in total** | **Tested adults** | **Number of tests** | **Incidence test rate (1000 persons/week)** | **Relative risk of test for SARS-CoV-2** *IRR (95% CI)* | |
| | | | | | **Crude*** | **Adjusted†** |
| Household without young children | 2,629,821 | 1,535,998 | 3,147,294 | 32.0 | 1 (ref.) | 1 (ref.) |
| Household with young children (any) | 449,915 | 293,857 | 599,874 | 35.6 | 1.07 (1.07,1.08) | 1.09 (1.09,1.09) |
| 1 | 341,132 | 222,927 | 454,937 | 35.6 | 1.07 (1.07,1.08) | 1.09 (1.08,1.09) |
| 2 | 104,125 | 68,103 | 139,232 | 35.7 | 1.08 (1.08,1.09) | 1.11 (1.10,1.11) |
| 3+ | 4,658 | 2,827 | 5,705 | 32.7 | 0.99 (0.96,1.01) | 1.04 (1.01,1.06) |

*Crude: Age and gender adjusted only.

†Adjusted: Furthermore adjusted for urbanicity, ethnicity, and comorbidities.
