## Supplementary material for "SARS-CoV-2 infection in households with and without young children: Nationwide cohort study": STROBE checklist

STROBE Statement—Checklist of items that should be included in reports of cohort studies for A. Husby et al. "**SARS-CoV-2 infection in households with and without young children: Nationwide cohort study**".

| | Item No | Recommendation | Description for manuscript |
|---|---|---|---|
| **Title and abstract** | 1 | (*a*) Indicate the study's design with a commonly used term in the title or the abstract | Reported in abstract |
| | | (*b*) Provide in the abstract an informative and balanced summary of what was done and what was found | Reported in abstract |
| **Introduction** | | | |
| Background/rationale | 2 | Explain the scientific background and rationale for the investigation being reported | Reported in introduction |
| Objectives | 3 | State specific objectives, including any prespecified hypotheses | Reported in introduction |
| **Methods** | | | |
| Study design | 4 | Present key elements of study design early in the paper | Reported introduction and methods |
| Setting | 5 | Describe the setting, locations, and relevant dates, including periods of recruitment, exposure, follow-up, and data collection | Reported in methods |
| Participants | 6 | (*a*) Give the eligibility criteria, and the sources and methods of selection of participants. Describe methods of follow-up | Reported in methods |
| | | (*b*) For matched studies, give matching criteria and number of exposed and unexposed | Not applicable |
| Variables | 7 | Clearly define all outcomes, exposures, predictors, potential confounders, and effect modifiers. Give diagnostic criteria, if applicable | Reported in methods |
| Data sources/ measurement | 8* | For each variable of interest, give sources of data and details of methods of assessment (measurement). Describe comparability of assessment methods if there is more than one group | Reported in methods |
| Bias | 9 | Describe any efforts to address potential sources of bias | Reported in methods and discussion |
| Study size | 10 | Explain how the study size was arrived at | Reported in in introduction and methods |
| Quantitative variables | 11 | Explain how quantitative variables were handled in the analyses. If applicable, describe which groupings were chosen and why | Reported in methods (statistical analysis) |
| Statistical methods | 12 | (*a*) Describe all statistical methods, including those used to control for confounding | Reported in methods (statistical analysis) |
| | | (*b*) Describe any methods used to examine subgroups and interactions | Reported in methods (statistical analysis) |
| | | (*c*) Explain how missing data were addressed | Reported in methods (statistical analysis) and Table 1 |

| | | | |
|---|---|---|---|
| | | (*d*) If applicable, explain how loss to follow-up was addressed | Reported in methods (statistical analysis) |
| | | (*e*) Describe any sensitivity analyses | Reported in methods, results, discussion, and supplement |
| **Results** | | | |
| Participants | 13* | (a) Report numbers of individuals at each stage of study—eg numbers potentially eligible, examined for eligibility, confirmed eligible, included in the study, completing follow-up, and analysed | Reported in results and Table 1 |
| | | (b) Give reasons for non-participation at each stage | Not applicable (nationwide longitudinal study) |
| | | (c) Consider use of a flow diagram | Not applicable |
| Descriptive data | 14* | (a) Give characteristics of study participants (eg demographic, clinical, social) and information on exposures and potential confounders | Reported in results and Table 1 |
| | | (b) Indicate number of participants with missing data for each variable of interest | Reported in Table 1 |
| | | (c) Summarise follow-up time (eg, average and total amount) | Reported in methods |
| Outcome data | 15* | Report numbers of outcome events or summary measures over time | Reported in results, Table 2 and Table 3 |
| Main results | 16 | (*a*) Give unadjusted estimates and, if applicable, confounder-adjusted estimates and their precision (eg, 95% confidence interval). Make clear which confounders were adjusted for and why they were included | Reported in Table 2 and Table 3 |
| | | (*b*) Report category boundaries when continuous variables were categorized | Not applicable |
| | | (*c*) If relevant, consider translating estimates of relative risk into absolute risk for a meaningful time period | Not applicable |
| Other analyses | 17 | Report other analyses done—eg analyses of subgroups and interactions, and sensitivity analyses | Reported in results and supplement |
| **Discussion** | | | |
| Key results | 18 | Summarise key results with reference to study objectives | Reported in discussion |
| Limitations | 19 | Discuss limitations of the study, taking into account sources of potential bias or imprecision. Discuss both direction and magnitude of any potential bias | Reported in discussion |
| Interpretation | 20 | Give a cautious overall interpretation of results considering objectives, limitations, multiplicity of analyses, results from similar studies, and other relevant evidence | Reported in discussion |
| Generalisability | 21 | Discuss the generalisability (external validity) of the study results | Reported in discussion |
| **Other information** | | | |

| | | | |
|---|---|---|---|
| Funding | 22 | Give the source of funding and the role of the funders for the present study and, if applicable, for the original study on which the present article is based | Not applicable. |

*Give information separately for exposed and unexposed groups.
